# A New psychosocial goal-setting and manualised support intervention for Independence in Dementia (NIDUS-Family) versus goal-setting and routine care: longer term outcomes of a single-masked, phase 3, superiority Randomised Controlled Trial

**DOI:** 10.1101/2025.03.05.25323403

**Authors:** Melisa Yilmaz, Victoria Vickerstaff, Jessica Budgett, Julie A. Barber, Claudia Cooper

**Affiliations:** Wolfson Institute of Population Health, Queen Mary University of London; Department of Statistical Science and Biostatistics Group within the NIHR UCLH Biomedical Research Centre; Priment Clinical Trials Unit, Research department of Primary Care and Population Health, University College London

## Abstract

**Background:** NIDUS-Family is a manualised intervention, deliverable by non-clinical facilitators which is clinically (on Goal Attainment Scaling - GAS) and cost effective over 12 months.

**Aims:** To evaluate whether goal setting plus NIDUS-Family was more effective than control (goal-setting and routine care) in supporting dyads’ (family carers and care recipients with dementia) attainment of personalised goals at 18 and 24 months; and participant perceived goal relevance over 24-months.

**Method:** A single-masked, randomised controlled trial recruiting dyads from community settings. Randomisation used a 2:1 ratio (intervention: control). NIDUS-Family is tailored to goals dyads set by selecting modules involving behavioural interventions, carer support, psychoeducation, communication, enablement and environmental adaptations. It involved 6-8 video-call/telephone sessions over 6 months, then follow-ups 2-3 monthly for 6 months. Our primary outcome was GAS at 18 and 24 months. Secondary outcomes assessed care recipient functioning, quality of life, time until care home admission or death, carer anxiety and depression. Primary analysis, a mixed-effects model, accounted for randomization group, study site, time, intervention-arm facilitator and repeated measurements.

**Results:** In 2020-21, 204 participants were randomised to intervention and 98 to control. 164 (54.3%) and 141(46.7%) dyads completed 18 and 24-month outcomes respectively. In the primary analysis, including 277 participants contributing 6-, 12-, 18- or 24-month outcomes, adjusted GAS mean differences (intervention–control) at 18 and 24-months were 11.78 (95% CI (Confidence Interval) 6.64,16.93) and 8.67 (3.31,14.02). Secondary outcome comparisons were not significant. The hazard ratio for dying or care home admission was 0.80 (0.45,1.42) (intervention v control); and 0.87 (0.41,1.82) and 0.59 (0.26,1.33) for death and care home admission respectively. Of baseline GAS goals, carers considered 436 (78.0%) relevant at 18 and 383 (78.5%) at 24 months.

**Conclusions:** The NIDUS-Family intervention improved personalised attainment of GAS goals that remained relevant for most dyads, over two years.

**Trial Registration Number: ISRCTN11425138**.

## Introduction

Most people with dementia wish to remain living at home for as long as possible, valuing the independence, safety and familiarity it provides (1,2). In the UK, approximately 700,000 unpaid family carers provide support to individuals with dementia in their homes, often expressing a willingness to do "whatever it takes" (1,3). However, without adequate support and strategies that balance the needs of people living with dementia and carers, care at home may break down, resulting in a sudden transition to a care home facility (4). The UK National Institute for Health and Care Excellence (NICE) guidelines stress the importance of offering psychosocial and environmental interventions to reduce stress, address behavioural and sleep disturbances with personalised strategies, and provide carer support (5). Nevertheless, there remains a significant gap between policy and real-world implementation, leaving carers and individuals with dementia with inadequate support (6).

Psychosocial interventions that are written down in a manual so they are delivered consistently (standardised), can be facilitated by trained, supervised staff without clinical qualifications, increasing access to evidence-based dementia care. Standardised therapies at first seem discordant with the “personalisation agenda”, which recognises that care is most effective if individually tailored. We co-designed, with Patient and Public Involvement (PPI), the New Intervention for Independence in Dementia Study – Family (NIDUS-Family) psychosocial support intervention to be fully manualised, modular so it can be tailored to individual goals, and delivered by facilitators without formal clinical training. It uses Goal Attainment Scaling (GAS), a structured outcome measure to set goals around what is most meaningful to carer and care recipient with dementia pairs (henceforth dyads). NIDUS-Family significantly improved goal attainment for dyads compared with goal setting and routine care, and was cost-effective over one year (7). In this paper, we aim to test whether NIDUS-Family improved dyads’ goal attainment compared to the control condition (goal setting and routine care) for up to two years. As this is the first time that GAS has been used as a trial measure beyond a year, we also asked participants about the relevance of baseline goals at 18 and 24-month follow-up.

## Methods

### Study design and participants

The authors assert that all procedures contributing to this work comply with the ethical standards of the relevant national and institutional committees on human experimentation and with the Helsinki Declaration of 1975, as revised in 2013. All procedures involving human subjects/patients were approved by Camden and King’s Cross Research Ethics Committee (19/LO/1667) on Jan 7, 2020. A substantial protocol amendment (approved Sept 19, 2022) added 18-month and 24-month follow-ups.

NIDUS-Family was a two-armed, parallel group, single masked, multi-site, superiority randomised controlled trial, for which the protocol (8) and 12-month primary clinical and cost-effectiveness outcomes are published (9,10).

NIDUS-family trial participant dyads were individuals with dementia and their informal (family or friend) carers (henceforth carers). Inclusion criteria were, for the person with dementia, a documented dementia diagnosis, regardless of type or severity, and living in their own home; and for carers, being in at least weekly face-to-face or telephone contact with the person with dementia and speaking English sufficiently to complete outcome measures (where the person with dementia did not speak English, we used interpreters to engage them as far as they were able during the intervention). We excluded dyads where the person with dementia was likely to be in their last six months of life, either member was participating in another study, or if the carer lacked the capacity to provide consent or could not identify a minimum of three appropriate GAS goals. Sex was self-reported. Written or verbal informed consent was obtained from all participants. Verbal consent was witnessed and formally recorded. Participant recruitment for the trial occurred between 30/04/2020-09/05/2021.

### Randomisation

The allocation process was managed via a remote web-based system provided by the PRIMENT Clinical Trials Unit (CTU) at University College London. Randomisation was blocked and stratified by site using a 2:1 allocation ratio (intervention: control). The randomisation status was concealed from researchers collecting outcome data from carers. It was not possible to blind participants or facilitators to their assigned group.

### Interventions/Procedures

Participants in the intervention arm received NIDUS-family, a manualised intervention that can be tailored to personal goals of people living with dementia and their families. It utilises components of behavioural management, carer support, psychoeducation, communication and coping skills training, enablement, and environmental adaptations, with modules selected to address dyads’ selected goals (11). NIDUS-Family was delivered by university-employed facilitators, without previous clinical training or clinical qualifications, who received a manualised training from the study team focusing on clinical skills and module delivery. Facilitators delivered 6-8 manualised sessions in the first 6 months, by video or telephone. These were tailored to participant GAS goals set at baseline (see outcome measures below). This was followed by 30-min catch-up telephone or video calls at 2–3-month intervals in the next 6 months, to review progress towards goals, implementation of action plans, and to troubleshoot difficulties following a standard guide.

Participants in the control condition received usual routine care and completed goal setting prior to randomisation.

### Outcome measures

The trial primary outcome was family carer-rated Goal Attainment Scaling (GAS), a valid and reliable measure that is responsive to change in function in people with dementia living at home (up to 12 months) and has been adapted to dementia (12,13). Trained researchers collaborated with carers and individuals living with dementia to establish three to five SMART goals (specific, measurable, achievable, realistic, and time-bound) tailored to domains such as cognition, daily living activities, self-care, mood, behaviour, and mobility. Goals were designed to support the person with dementia in living well or maintaining independence at home over the following year, provided they aligned with the intervention’s scope. While goals could address carer wellbeing or support if these influenced the person’s functioning or quality of life, at least one goal was required to directly focus on the individual with dementia. At baseline, family carers set criteria for how to evaluate ‘performance’ on goals set, on a 5-point scale ranging from “much worse” to “much better” than expected. They were then reminded of these criteria at follow-ups and asked to rate goal performance. As people had different goals and numbers of goals, a summary formula standardised the degree of goal attainment, analysed as a change score. Researchers also independently rated participant GAS attainment after the main outcome battery was complete and based on their conversations. They sought to record this independently of the carer rating by recording it ahead of the carer rating. The primary outcome in this study was family-carer rated GAS scores at 18 and 24 months. At the 18- and 24-month follow-ups, dyads were asked ‘is this goal still relevant to you / the person you care for?’ and the response was recorded as a yes or no. If a dyad reported that their goal was no longer relevant, they were asked why this was the case.

Information about the living status of the person with dementia was obtained at 18 and 24 months, including whether they were still alive (and if not, the date of death), whether they had moved to a care home, whether the move was permanent, the date of care home admission, and the length of stay in days. To minimise assessment burden, other outcomes were only completed at 18 months. These were: the Disability Assessment for Dementia scale, a measure of functional independence (basic and instrumental activities of daily living)(12); DEMQOL, a measure of quality of life of people with dementia, completed by family carers (DEMQOL-proxy) and people with dementia if they were able (DEMQOL) (14), the Hospital Anxiety and Depression Scale to measure family carers psychological morbidity (15) and the modified Client Service Receipt Inventory (CSRI) (16).

### Statistical analysis

Intervention effects at 18 and 24 months were estimated as differences in mean family carer-rated GAS scores between allocation groups, obtained from a three-level mixed-effects model using all available repeated measures at 6, 12, 18, and 24 months. The model included random effects for intervention-arm facilitator and the repeated measurements, with fixed effects for randomization group, study site, and time point. An interaction term between randomization group and time point was included to estimate intervention effects at each follow-up point. We used similar models to estimate treatment effects for the secondary DemQol Proxy and researcher-rated GAS outcomes. The model for DemQol-proxy additionally included adjustment for baseline DemQol-proxy score. Other outcome scores were summarised by group.

Time to permanent care home admission or death was calculated relative to baseline and summarised by randomised group using Kaplan-Meier plots (Figure 1S and 2S, Supplementary material) for each event type and for the composite (death or care home transition). To formally compare groups the time to admission/death was analysed using a parametric shared frailty model, assuming a Weibull survival model, allowing for intervention arm facilitator clustering and adjusting for study site. Those lost or withdrawn before death or care home admission were censored at their last follow-up point. We also fitted a competing risks model, adjusted for study site, to obtain separate effect estimates for death and care home transition.

**Figure 1:**
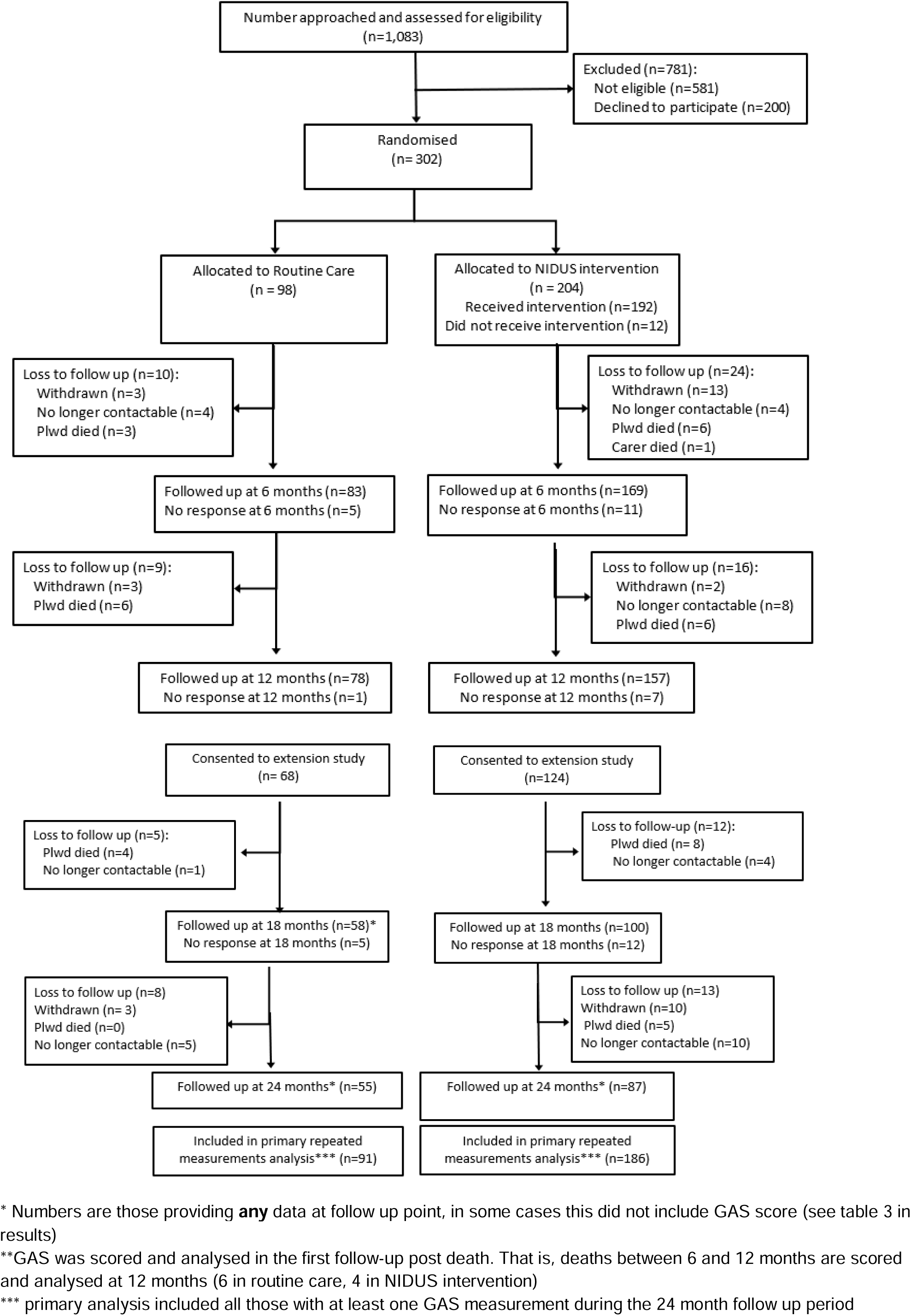
Consort diagram for NIDUS-family trial.

Analyses were intention to treat and included all available data, assuming any missing values were missing at random. For our primary outcome we conducted sensitivity analyses to consider the impact of missing data on our results. We refitted our main model based on datasets completed using multiple imputation (MI); our imputation models included repeated outcome measurements, socio-demographic baseline data, and other variables potentially related to missingness and outcome (DemQol proxy baseline score and DAD baseline score), with imputations performed by study arm. A pattern-mixture approach was used to investigate missing not at random scenarios. A range of δ values (0, 4, 8, 12) was subtracted from the MI GAS scores, and regression models with fixed effects for site and treatment group were fitted for the 18- and 24-month outcomes. In addition, a worst-case sensitivity analysis was conducted where participants admitted to a care home (permanently) or who had died had their missing GAS goal values imputed with a value of -2, indicating “much worse” than expected.

At 18 and 24 months, we asked participants for each goal set, whether it still felt relevant. We conducted a content analysis of free text responses to identify the main reasons why goals were no longer considered to be relevant, using an inductive content analysis approach (17,18).

## Results

### Sample description

From 302 dyads randomised, the primary outcome was available for 164 (54.3%) at 18-month follow up, and 141 (46.7%) at 24-months. Figure 1 (Consort diagram) illustrates the flow of participants through the study.

Tables 1 and 2 show the baseline characteristics of people living with dementia and family carers.

**Table 1:** Baseline participant characteristics by arm.

|  | Randomised group |  | NIDUS-Family |  | Total |  |
| --- | --- | --- | --- | --- | --- | --- |
|  | Routine care<br>(n=98) |  | intervention<br>(n=204) |  | (n=302) |  |
| Age | 80.3 | (8.7) | 79.7 | (8.0) | 79.9 | (8.2) |
| Ethnicity |  |  |  |  |  |  |
| White | 87 | (88.8) | 179 | (87.7) | 266 | (88.1) |
| Mixed | 2 | (2.0) | 2 | (1.0) | 4 | (1.3) |
| Asian | 5 | (5.1) | 12 | (5.9) | 17 | (5.6) |
| Black | 2 | (2.0) | 9 | (4.4) | 11 | (3.6) |
| Other | 2 | (2.0) | 2 | (1.0) | 4 | (1.3) |
| First language |  |  |  |  |  |  |
| English | 83 | (84.7) | 177 | (86.8) | 260 | (86.1) |
| Other | 15 | (15.3) | 27 | (13.2) | 42 | (13.9) |
| Gender |  |  |  |  |  |  |
| Male | 38 | (38.8) | 95 | (46.6) | 133 | (44.0) |
| Female | 60 | (61.2) | 109 | (53.4) | 169 | (56.0) |
| Marital status |  |  |  |  |  |  |
| Married/civil partnership | 57 | (58.2) | 116 | (56.9) | 173 | (57.3) |
| Divorced | 7 | (7.1) | 10 | (4.9) | 17 | (5.6) |
| Single | 0 | (0.0) | 5 | (2.5) | 5 | (1.7) |
| Co-habiting | 1 | (1.0) | 4 | (2.0) | 5 | (1.7) |
| Widowed | 33 | (33.7) | 67 | (32.8) | 100 | (33.1) |
| Other | 0 | (0.0) | 2 | (1.0) | 2 | (0.7) |
| Education (n=296) |  |  |  |  |  |  |
| Higher degree | 9 | (9.2) | 24 | (12.1) | 33 | (11.1) |
| Degree | 18 | (18.4) | 38 | (19.2) | 56 | (18.9) |
| A level (or equivalent) | 9 | (9.2) | 16 | (8.1) | 25 | (8.4) |
| HNC/HND (or equivalent) | 7 | (7.1) | 14 | (7.1) | 21 | (7.1) |
| NVQ (or equivalent) | 5 | (5.1) | 7 | (3.5) | 12 | (4.1) |
| GCSE (or equivalent) | 17 | (17.3) | 30 | (15.2) | 47 | (15.9) |
| School Leaving Certificate | 16 | (16.3) | 39 | (19.7) | 55 | (18.6) |
| no formal qualifications | 17 | (17.3) | 30 | (15.2) | 47 | (15.9) |
| Living situation |  |  |  |  |  |  |
| Live alone | 22 | (22.4) | 62 | (30.4) | 84 | (27.8) |
| Live with partner/spouse | 52 | (53.1) | 107 | (52.5) | 159 | (52.6) |
| Live with children | 16 | (16.3) | 23 | (11.3) | 39 | (12.9) |
| Other | 8 | (8.2) | 12 | (5.9) | 20 | (6.6) |
| Accommodation |  |  |  |  |  |  |
| Council rented | 5 | (5.1) | 15 | (7.4) | 20 | (6.6) |
| Housing association rented | 5 | (5.1) | 9 | (4.4) | 14 | (4.6) |
| Private rented | 3 | (3.1) | 10 | (4.9) | 13 | (4.3) |
| Owner-occupied | 82 | (83.7) | 155 | (76.0) | 237 | (78.5) |
| Other | 3 | (3.1) | 15 | (7.4) | 18 | (6.0) |
| Dementia diagnosis |  |  |  |  |  |  |
| Alzheimer's Disease | 44 | (44.9) | 95 | (46.6) | 139 | (46.0) |
| Vascular dementia | 10 | (10.2) | 28 | (13.7) | 38 | (12.6) |
| Lewy body dementia | 3 | (3.1) | 7 | (3.4) | 10 | (3.3) |
| Frontotemporal dementia | 2 | (2.0) | 6 | (2.9) | 8 | (2.6) |
| Other | 26 | (26.5) | 58 | (28.4) | 84 | (27.8) |
| Unable to specify | 13 | (13.3) | 10 | (4.9) | 23 | (7.6) |

**Table 2:**
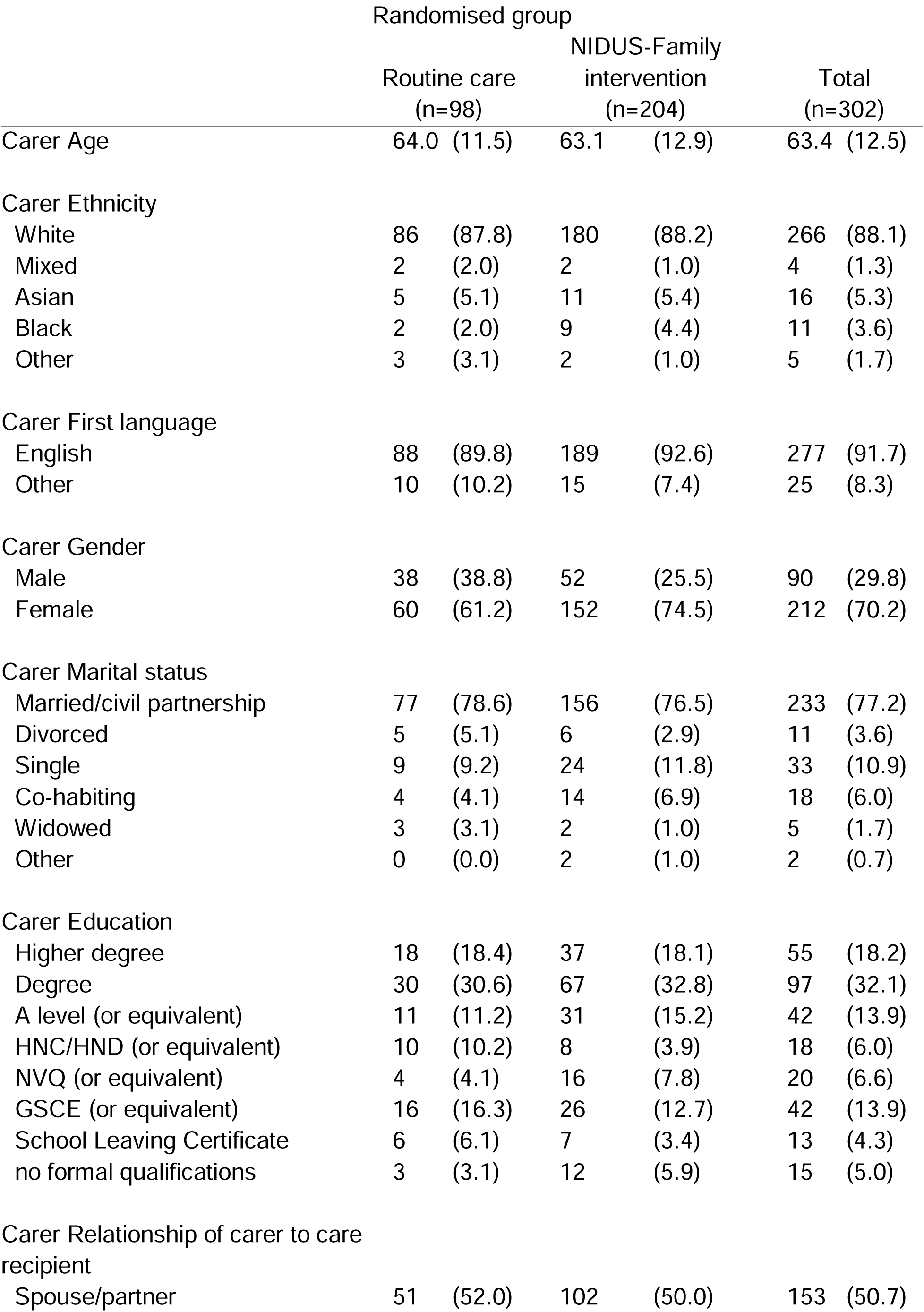
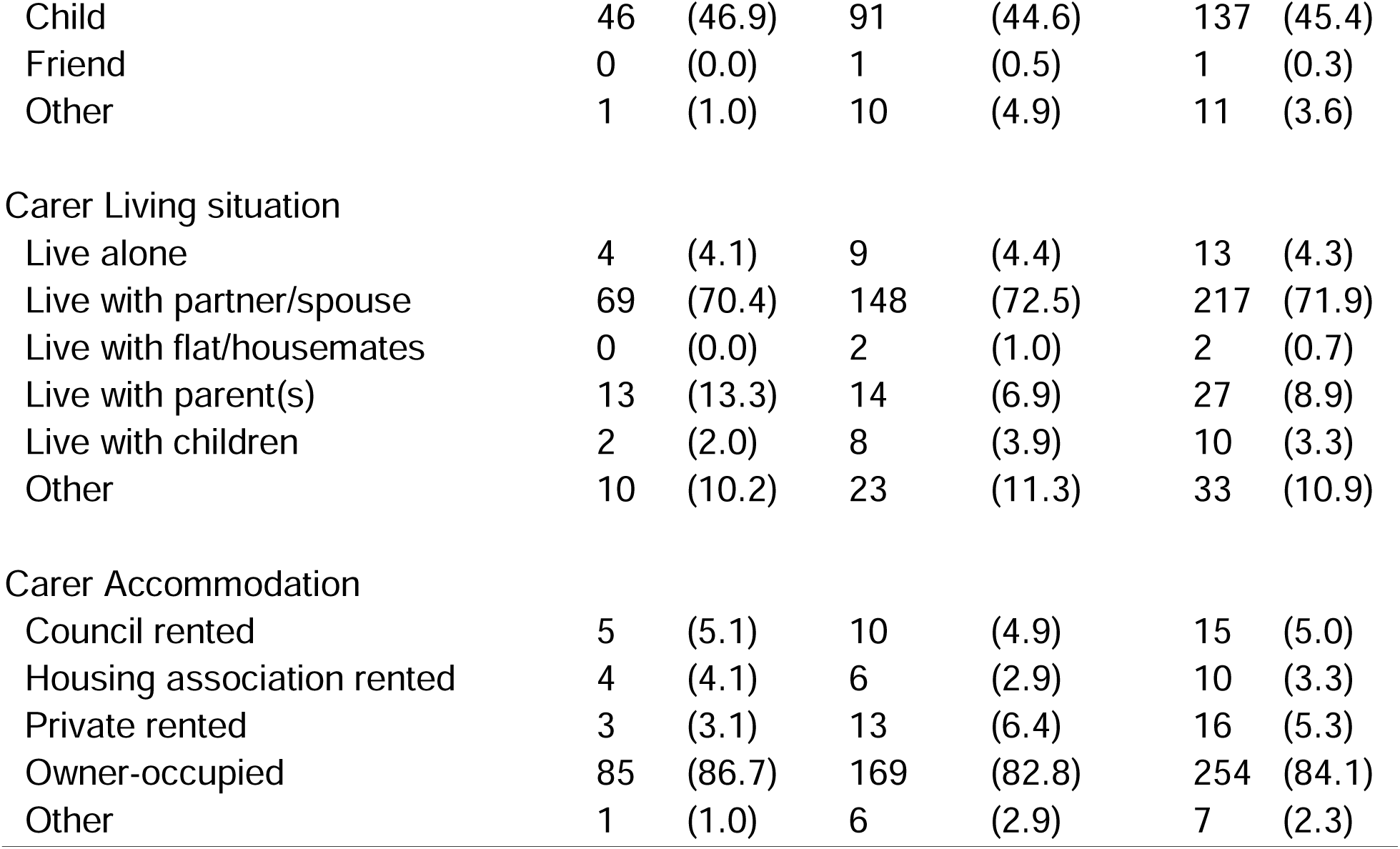
Carer characteristics by arm.

Tables 1S-4S (Supplemental material) show the characteristics of people living with dementia and family carers who did and did not take part in the extension study. Compared to those who did not take part, people with dementia and carers who took part in the extension study were slightly younger, but otherwise characteristics were very similar.

### Comparison of primary outcome between arms

The mean family carer-rated GAS score was significantly higher in the intervention than the control arm at all time points (Figure 2).

**Figure 2:**
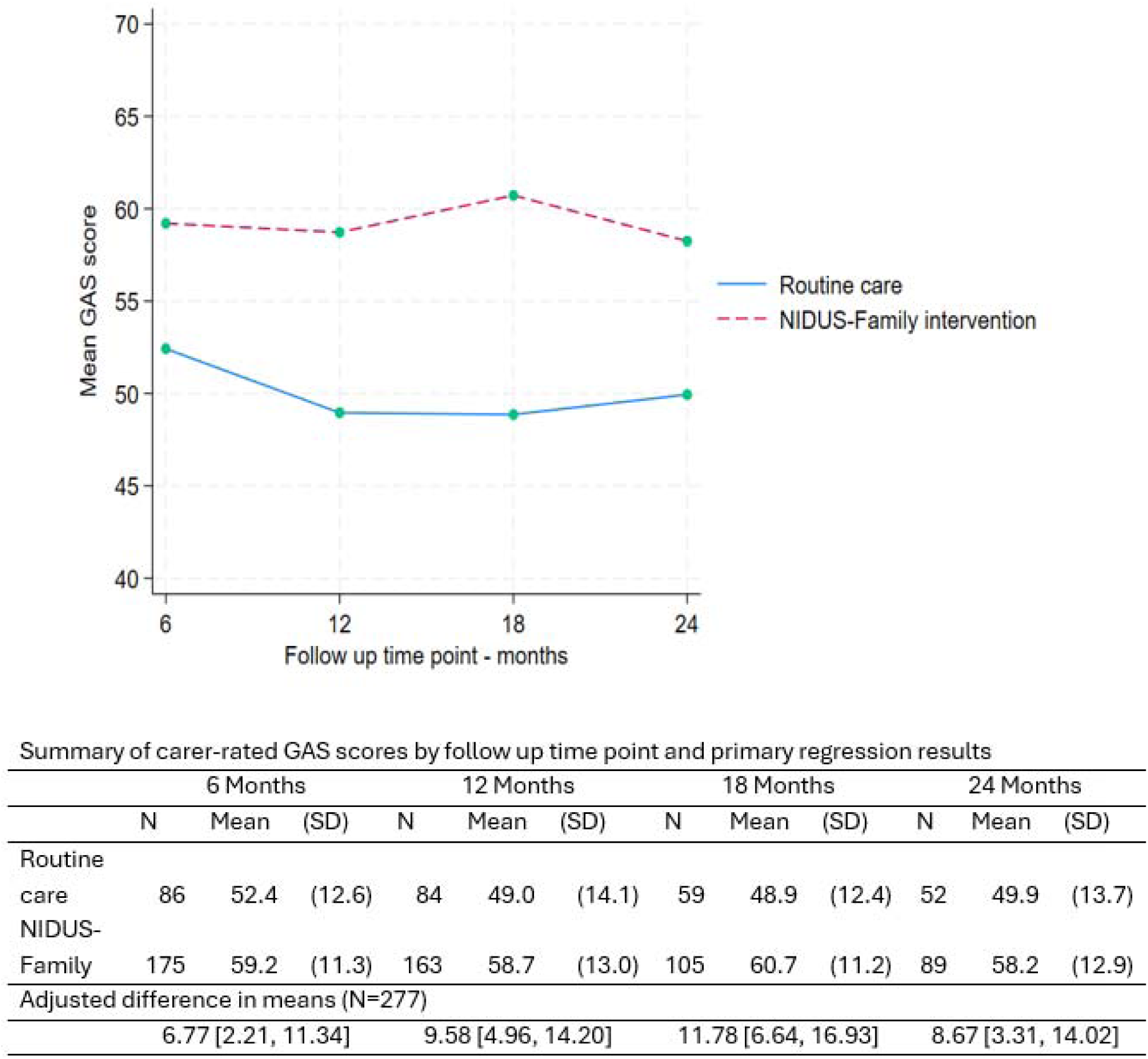
The primary outcome (carer-rated GAS scores) over 24 months, by arm.

We included 277 people living with dementia who had at least one primary outcome measurement recorded (at 6, 12, 18, or 24 months) in our mixed effects model. In this model, the adjusted difference in means (NIDUS – control group) at 18-month follow-up was 11.78 (95% CI 6.64,16.93), and 8.67 (95% CI 3.31,14.02) at 24 months. These estimates were very similar in multiple imputation and worse-case scenario sensitivity analyses (Table 5S). Table 5S shows findings from the pattern-mixture approach we used to investigate missing not at random scenarios. For the 18-month outcomes, the intervention effect remained significant with all assumptions; while for 24-month outcomes, it remained significant for all but some of the most extreme scenarios.

### Comparison of secondary outcomes

Table 3 summarises secondary outcome scores at all time points.

**Table 3:**
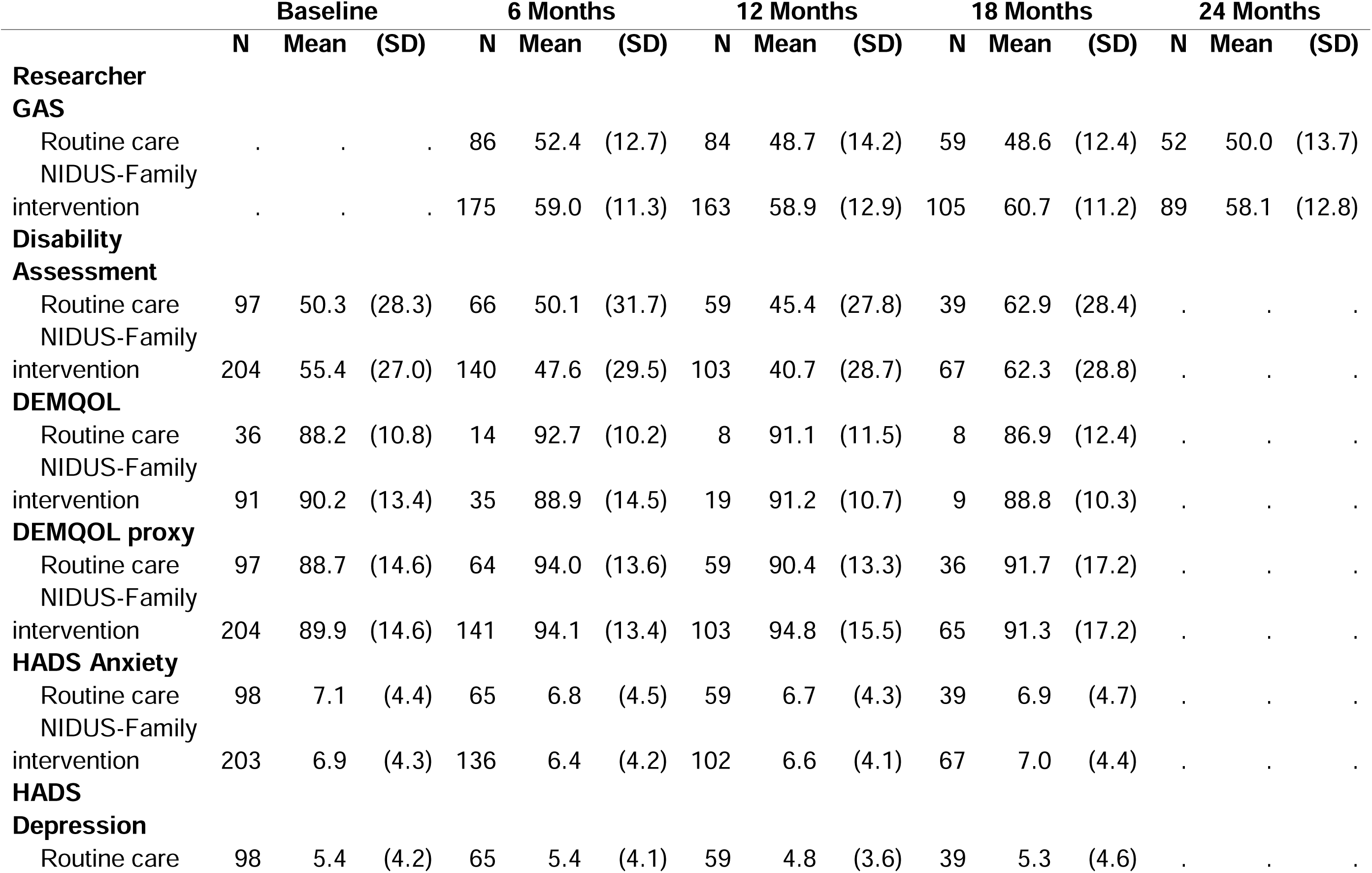
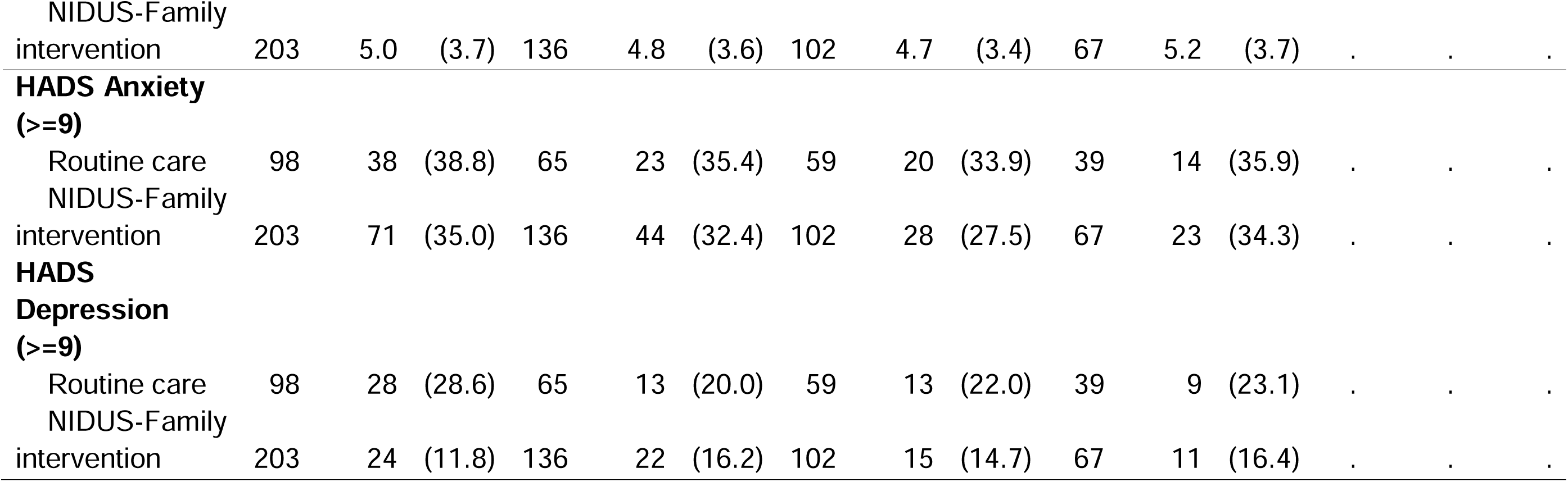
Summaries of secondary outcome scores at each follow up point, by arm.

Researcher-rated GAS mean scores were higher in the NIDUS-Family intervention compared to the control arm across all time points. Differences in means between groups were significant in adjusted analyses; 11.99 (95% CI: 8.09, 15.90) at 18 months and 8.94 (95% CI: 4.81, 13.06) at 24 months. The adjusted treatment effect estimates for DEMQOL-proxy were not statistically significant; adjusted difference in means of 2.36 (95% CI: -1.59, 6.30) at 18 months. Treatment effect estimates for these outcomes are reported in Table 6S (supplementary material). Other scores were similar across arms (Table 3).

### Care home admission and mortality (Table 7S)

By 24-months, 15 (15.3%) control arm participants and 25 (12.2%) intervention arm participants were know to have moved to a care home; while 13 (13.3%) control arm and 25 (12.3%) intervention arm participants were known to have died. The hazard ratio for dying or moving to a care home was 0.80 (95% CI: 0.45, 1.42) for the NIDUS-family intervention compared to routine care; Sub Hazard Ratios for death and care home admission were 0.87 (95% CI: 0.41, 1.82), and 0.59 (95% CI: 0.26, 1.33) respectively. The latter indicates a reduction in the hazard of permanent care home admission of 41% in the intervention relative to control group, although this is not statistically significant.

### Goal Relevance

The 164 carers rating GAS goals at 18 months evaluated performance on 559 goals, of which 436 (78.0%) were reported by carers as still feeling relevant. The 141 carers rating GAS goals at 24 months evaluated performance on 488 goals, of which 383 (78.5%) still felt relevant.

The reasons why goals no longer felt relevant included: the goals being achieved (7 goals at 18 months, 12 goals at 24 months); the person with dementia having passed away (30 goals at 18 months, 4 goals at 24 months); moving to a care home (15 goals at 18 months, 9 goals at 24 months); the goals becoming unrealistic due to a decline of the person living with dementia’s health (12 goals at 18 months, 30 goals at 24 months); the goals no longer aligning with the dyad’s priorities, as they had accepted the person’s current functional state and shifted focus (5 goals at 18 months, 14 goals at 24 months); no observable change (4 goals at 18 months, 1 goal at 24 months); or external circumstances or stressors preventing carers to progress towards the goals (3 goals at 24 months), as detailed in Table 8S (supplementary material). In Table 9S (supplementary material), we provide examples for each of these reasons for goals no longer feeling relevant.

## Discussion

We have previously reported that goal-setting and NIDUS-Family improved dyads’ goal attainment, compared to goal-setting and routine care, and was cost-effective over one year (9,10). In the current study, we reported that it continued to improve goal attainment for up to two years.

The reduction in the rate of care home admission we found in the intervention group was not statistically significant, but was of a similar magnitude to findings previously reporting that good quality support can reduce care home admissions among people with dementia by around a third. In a meta-analysis (n=9053) the odds ratio (OR) for institutionalisation in interventions designed to improve home support versus control was 0.66, (95% confidence interval (CI) 0.43–0.99). In two US studies which informed the NIDUS-family theoretical development (19), home-support interventions for family carers and people living with dementia also delayed institutionalisation by around one third (HRs 0.65, 95% confidence interval [CI], 0.45 to 0.94 (20); and 0.63, 0.42 to 0.94 (21)). A commonality of these interventions was a goal-focused approach to supporting care recipients and family carers with dementia. This strategy, of identifying key factors dyads prioritise to enable continued care at home may be critical to extending time lived at home. There is little evidence that carer psychological stress alone increases institutionalisation (22), or that interventions focused primarily on reducing carer stress prevent it (23).

This is to our knowledge the first use of GAS as a trial outcome measure over a period greater than a year, with most studies utilising it over three months or less (24). It is encouraging that nearly eight in ten goals set still felt relevant for up to two years, suggesting that GAS can be a useful outcome over this longer period. Average proxy-rated quality of life scores in the intervention versus control arm were not statistically significant, and of a similar magnitude to those recorded at 12 months in the NIDUS-family trial; we previously noted that this may be a clinically important effect size (25) that we lacked the power to detect. GAS is a measure that is very responsive to change. It may also be measuring a conceptually different construct to life quality; we asked dyads to set goals towards living as long and as well as possible at home; many people with dementia and family carers consider remaining at home a priority, even where it brings challenges.

There are some limitations to our research. We asked carers to set goals, to which people living with dementia contributed to the extent they were able. We cannot independently verify how goals reflected the likely wishes of people with dementia who lacked capacity. We did not include people with dementia without a regular carer, or whose carer could access the intervention written in English. Participants originally consented to complete outcomes for a year, and while most agreed to take part in this extension study, response rates were lower than for previous time points. We maximised participation by minimising assessment burden and thus had very limited measures at 24 months. Our sensitivity and multiple imputation models have evaluated the potential impact of missing data however and indicate that our findings are robust.

NIDUS-family is delivered by professionals without formal clinical training, so can extend the workforce capable of delivering evidence-based dementia care. Nonetheless, sufficient workforce resources are needed, and limited resources were noted to be a significant implementation barrier to wider delivery across health and social care sectors in our recent pre-implementation study (26). Major cuts in social care budgets increase the risk of high-cost care home admissions which older people do not want. (27). In England, inadequate adult social care availability (for care home placements and home care support) has led to more requests for help been turned down in 2023/4, delaying hospital discharge, and reducing the capacity of people to live as independently as possible in their own homes (28). Limited dementia skills and training within the social care workforce is a further barrier to implementation (29).

The NIDUS-Family dementia care intervention improved personalised goal attainment, on GAS goals that remained relevant for most dyads, over two years. Most people with dementia prefer to remain in their own homes, and this is best enabled through providing good quality, personalised community health and social care. NIDUS-family is a potentially important tool to enable this, with materials available free of charge (thenidussstudy.co.uk), and deliverable after appropriate training and with supervision.

## Author contribution

All authors were involved in designing the study, carrying it out, analysing the data and writing the article.

## Transparency declaration

Claudia Cooper as guarantor and Julie Barber as senior statistician affirm that the manuscript is an honest, accurate, and transparent account of the study being reported; that no important aspects of the study have been omitted; and that any discrepancies from the study as planned (and, if relevant, registered) have been explained.

## Data sharing

Data collected for the study, including the statistical analysis plan, deidentified participant data and a data dictionary defining each field in the set, will be made available to others on receipt by Priment CTU of a reasonable request, at any date after publication of this paper. All requests will be reviewed by Priment CTU in line with Priment CTU guidance on sharing data and anonymising data. This process is to ensure that the request is reasonable and the data set is suitably anonymised. The study protocol is available open access. Intervention materials are available without cost, subject to a CC BY-NC-ND license held by Claudia Cooper, Chief Investigator.

## Data Availability

Data collected for the study, including the statistical analysis plan, deidentified participant data and a data dictionary defining each field in the set, will be made available to others on receipt by Priment CTU of a reasonable request, at any date after publication of this paper. All requests will be reviewed by Priment CTU in line with Priment CTU guidance on sharing data and anonymising data.

## References

1. Rapaport P, Burton A, Leverton M, Herat-Gunaratne R, Beresford-Dent J, Lord K, et al. “I just keep thinking that I don’t want to rely on people.” a qualitative study of how people living with dementia achieve and maintain independence at home: stakeholder perspectives. BMC Geriatr. 2020 Jan 3;20(1):5.

2. Herat-Gunaratne R, Cooper C, Mukadam N, Rapaport P, Leverton M, Higgs P, et al. ‘In the Bengali Vocabulary, There Is No Such Word as Care Home’: Caring Experiences of UK Bangladeshi and Indian Family Carers of People Living With Dementia at Home. The Gerontologist. 2020 Feb;60(2):331–9.

3. Alzheimer’s Society. Carers UK’s ‘State of Caring 2021’ report – Alzheimer’s Society responds | Alzheimer’s Society [Internet]. 2021 [cited 2024 Dec 6]. Available from: https://www.alzheimers.org.uk/news/2024-11-22/carers-uks-state-caring-2021-report-alzheimers-society-responds

4. Lord K, Livingston G, Robertson S, Cooper C. How people with dementia and their families decide about moving to a care home and support their needs: development of a decision aid, a qualitative study. BMC Geriatr. 2016;16(1):68.

5. NICE. Recommendations | Dementia: assessment, management and support for people living with dementia and their carers | Guidance | NICE [Internet]. NICE; 2018 [cited 2024 Nov 21]. Available from: https://www.nice.org.uk/guidance/ng97/chapter/Recommendations

6. Seidel K, Quasdorf T, Haberstroh J, Thyrian JR. Adapting a Dementia Care Management Intervention for Regional Implementation: A Theory-Based Participatory Barrier Analysis. Int J Environ Res Public Health. 2022 Jan;19(9):5478.

7. Isaaq A, Cooper C, Vickerstaff V, Barber JA, Walters K, Lang IA, et al. Cost-utility of a new psychosocial goal-setting and manualised support intervention for independence in dementia (NIDUS-Family) versus goal setting and routine care: an economic evaluation embedded within a randomised controlled trial. Lancet Healthy Longev. 2025 Feb 17;100676.

8. Burton A, Rapaport P, Palomo M, Lord K, Budgett J, Barber J, et al. Clinical and cost-effectiveness of a New psychosocial intervention to support Independence in Dementia (NIDUS-family) for family carers and people living with dementia in their own homes: a randomised controlled trial. Trials [Internet]. 2021 Dec 2;22(1). Available from: 10.1186/s13063-021-05851-z

9. Cooper C, Vickerstaff V, Barber J, Phillips R, Ogden M, Walters K, et al. A psychosocial goal-setting and manualised support intervention for independence in dementia (NIDUS-Family) versus goal setting and routine care: a single-masked, phase 3, superiority, randomised controlled trial. Lancet Healthy Longev. 2024 Feb;5(2):e141–51.

10. Isaaq A, Cooper C, Vickerstaff V, Barber J, Walters K, Lang I, et al. Cost-utility of a new psychosocial goal-setting and manualised support intervention for Independence in Dementia (NIDUS-Family) versus goal-setting and routine care: economic evaluation embedded within a randomised controlled trial. [Internet]. medRxiv; 2024 [cited 2024 Aug 26]. p. 2024.08.24.24312530. Available from: https://www.medrxiv.org/content/10.1101/2024.08.24.24312530v1

11. Wyman D, Butler LT, Morgan-Trimmer S, Bright P, Barber J, Budgett J, et al. Process evaluation of a New psychosocial goal-setting and manualised support intervention for Independence in Dementia (NIDUS-Family). Age Ageing. 2024 Aug 6;53(8):afae181.

12. Feldman H, Sauter A, Donald A, Gélinas I, Gauthier S, Torfs K, et al. The disability assessment for dementia scale: a 12-month study of functional ability in mild to moderate severity Alzheimer disease. Alzheimer Dis Assoc Disord. 2001;15(2):89– 95.

13. Rockwood K, Graham JE, Fay S, ACADIE Investigators. Goal setting and attainment in Alzheimer’s disease patients treated with donepezil. J Neurol Neurosurg Psychiatry. 2002 Nov;73(5):500–7.

14. Smith SC, Lamping DL, Banerjee S, Harwood R, Foley B, Smith P, et al. Measurement of health-related quality of life for people with dementia: development of a new instrument (DEMQOL) and an evaluation of current methodology. Health Technol Assess Winch Engl. 2005 Mar;9(10):1–93, iii–iv.

15. Zigmond AS, Snaith RP. The hospital anxiety and depression scale. Acta Psychiatr Scand. 1983 Jun;67(6):361–70.

16. Beecham J, Knapp M. Costing psychiatric interventions. In: Measuring mental health needs. London, England: Gaskell/Royal College of Psychiatrists; 1992. p. 163–83.

17. Budgett J, Sommerlad A, Kupeli N, Zabihi S, Rockwood K, Cooper C. Personalized goals of people living with dementia and family carers: A content analysis of goals set within an individually tailored psychosocial intervention trial. Alzheimers Dement Transl Res Clin Interv. 2024 Jul;10(3):e12493.

18. Elo S, Kyngäs H. The qualitative content analysis process. J Adv Nurs. 2008 Apr;62(1):107–15.

19. Lord K, Beresford-Dent J, Rapaport P, Burton A, Leverton M, Walters K, et al. Developing the New Interventions for independence in Dementia Study (NIDUS) theoretical model for supporting people to live well with dementia at home for longer: a systematic review of theoretical models and Randomised Controlled Trial evidence. Soc Psychiatry Psychiatr Epidemiol. 2020;55(1):1–14.

20. Mittelman MS, Haley WE, Clay OJ, Roth DL. Improving caregiver well-being delays nursing home placement of patients with Alzheimer disease. Neurology. 2006 Nov 14;67(9):1592–9.

21. Samus QM, Johnston D, Black BS, Hess E, Lyman C, Vavilikolanu A, et al. A multidimensional home-based care coordination intervention for elders with memory disorders: the maximizing independence at home (MIND) pilot randomized trial. Am J Geriatr Psychiatry Off J Am Assoc Geriatr Psychiatry. 2014 Apr;22(4):398–414.

22. Donnelly NA, Hickey A, Burns A, Murphy P, Doyle F. Systematic Review and Meta-Analysis of the Impact of Carer Stress on Subsequent Institutionalisation of Community-Dwelling Older People. PLOS ONE. 2015 Jun 2;10(6):e0128213.

23. Livingston G, Manela M, O’Keeffe A, Rapaport P, Cooper C, Knapp M, et al. Clinical effectiveness of the START (STrAtegies for RelaTives) psychological intervention for family carers and the effects on the cost of care for people with dementia: 6-year follow-up of a randomised controlled trial. Br J Psychiatry. 2020 Jan 1;1–8.

24. Logan B, Jegatheesan D, Viecelli A, Pascoe E, Hubbard R. Goal attainment scaling as an outcome measure for randomised controlled trials: a scoping review. BMJ Open. 2022 Jul 1;12(7):e063061.

25. Ballard C, Corbett A, Orrell M, Williams G, Moniz-Cook E, Romeo R, et al. Impact of person-centred care training and person-centred activities on quality of life, agitation, and antipsychotic use in people with dementia living in nursing homes: A cluster-randomised controlled trial. PLoS Med. 2018 Feb 6;15(2):e1002500.

26. Dar A, Budgett J, Zabihi S, Whitfield E, Lang I, Rapaport P, et al. Pre-implementation planning for a new personalised, dementia post-diagnostic support intervention: exploring the perspective of professional stakeholders. BJPsych Open. 2024 Sep;10(5):e139.

27. Knapp M, Chua KC, Broadbent M, Chang CK, Fernandez JL, Milea D, et al. Predictors of care home and hospital admissions and their costs for older people with Alzheimer’s disease: findings from a large London case register. BMJ Open. 2016 Nov 1;6(11):e013591.

28. Adult social care - Care Quality Commission [Internet]. [cited 2024 Dec 22]. Available from: https://www.cqc.org.uk/publications/major-report/state-care/2023-2024/access/asc

29. Delray S, Banerjee S, Zabihi S, Walpert M, Harrison-Dening K, Kenten C, et al. Systematic policy and evidence review to consider how dementia education and training is best delivered in the social care workforce, and how policy does or can enable its implementation in England [Internet]. medRxiv; 2024 [cited 2024 Aug 26]. p. 2024.08.24.24312532. Available from: https://www.medrxiv.org/content/10.1101/2024.08.24.24312532v1

