## Supplementary Materials for "A New psychosocial goal-setting and manualised support intervention for Independence in Dementia (NIDUS-Family) versus goal-setting and routine care: longer term outcomes of a single-masked, phase 3, superiority Randomised Controlled Trial"

**Table 1S: Baseline characteristics for people living with dementia participating in the extension study, by arm**

|  | **Randomised group** | | | | | | | | |
| --- | --- | --- | --- | --- | --- | --- | --- | --- | --- |
|  | **Routine care**  **N=68** | | | **NIDUS-Family intervention**  **N=124** | | | **Total**  **N=192** | | |
| **Age (years), mean (SD)** |  | 79.8 | (8.5) |  | 78.9 | (8.4) |  | 79.2 | (8.4) |
| **Ethnicity, n(%)** |  |  |  |  |  |  |  |  |  |
| White |  | 56 | (82.4) |  | 98 | (79.0) |  | 154 | (80.2) |
| White other |  | 6 | (8.8) |  | 14 | (11.3) |  | 20 | (10.4) |
| Mixed |  | 2 | (2.9) |  | 0 | (0.0) |  | 2 | (1.0) |
| Asian |  | 1 | (1.5) |  | 6 | (4.8) |  | 7 | (3.6) |
| Black |  | 1 | (1.5) |  | 4 | (3.2) |  | 5 | (2.6) |
| Other |  | 2 | (2.9) |  | 2 | (1.6) |  | 4 | (2.1) |
| **First language, n(%)** |  |  |  |  |  |  |  |  |  |
| English |  | 59 | (86.8) |  | 106 | (85.5) |  | 165 | (85.9) |
| Other |  | 9 | (13.2) |  | 18 | (14.5) |  | 27 | (14.1) |
| **Gender, n(%)** |  |  |  |  |  |  |  |  |  |
| Male |  | 28 | (41.2) |  | 57 | (46.0) |  | 85 | (44.3) |
| Female |  | 40 | (58.8) |  | 67 | (54.0) |  | 107 | (55.7) |
| **Marital status, n(%)** |  |  |  |  |  |  |  |  |  |
| Married/civil partnership |  | 41 | (60.3) |  | 68 | (54.8) |  | 109 | (56.8) |
| Divorced |  | 5 | (7.4) |  | 4 | (3.2) |  | 9 | (4.7) |
| Single |  | 0 | (0.0) |  | 3 | (2.4) |  | 3 | (1.6) |
| Co-habiting |  | 1 | (1.5) |  | 3 | (2.4) |  | 4 | (2.1) |
| Widowed |  | 21 | (30.9) |  | 44 | (35.5) |  | 65 | (33.9) |
| Other |  | 0 | (0.0) |  | 2 | (1.6) |  | 2 | (1.0) |
| **Education, n(%) (n=189)** |  |  |  |  |  |  |  |  |  |
| Higher degree |  | 7 | (10.3) |  | 13 | (10.7) |  | 20 | (10.6) |
| Degree |  | 12 | (17.6) |  | 23 | (19.0) |  | 35 | (18.5) |
| A level (or equivalent) |  | 7 | (10.3) |  | 11 | (9.1) |  | 18 | (9.5) |
| HNC/HND (or equivalent) |  | 6 | (8.8) |  | 10 | (8.3) |  | 16 | (8.5) |
| NVQ (or equivalent) |  | 4 | (5.9) |  | 6 | (5.0) |  | 10 | (5.3) |
| GCSE (or equivalent) |  | 12 | (17.6) |  | 22 | (18.2) |  | 34 | (18.0) |
| School Leaving Certificate |  | 9 | (13.2) |  | 21 | (17.4) |  | 30 | (15.9) |
| no formal qualifications |  | 11 | (16.2) |  | 15 | (12.4) |  | 26 | (13.8) |
| **Living situation, n(%)** |  |  |  |  |  |  |  |  |  |
| Live alone |  | 16 | (23.5) |  | 42 | (33.9) |  | 58 | (30.2) |
| Live with partner/spouse |  | 38 | (55.9) |  | 62 | (50.0) |  | 100 | (52.1) |
| Live with children |  | 9 | (13.2) |  | 12 | (9.7) |  | 21 | (10.9) |
| Other |  | 5 | (7.4) |  | 8 | (6.5) |  | 13 | (6.8) |
| **Accommodation, n(%)** |  |  |  |  |  |  |  |  |  |
| Council rented |  | 4 | (5.9) |  | 8 | (6.5) |  | 12 | (6.2) |
| Housing association rented |  | 4 | (5.9) |  | 6 | (4.8) |  | 10 | (5.2) |
| Private rented |  | 3 | (4.4) |  | 5 | (4.0) |  | 8 | (4.2) |
| Owner-occupied |  | 55 | (80.9) |  | 94 | (75.8) |  | 149 | (77.6) |
| Other |  | 2 | (2.9) |  | 11 | (8.9) |  | 13 | (6.8) |
| **Dementia diagnosis, n(%)** |  |  |  |  |  |  |  |  |  |
| Alzheimer’s Disease |  | 32 | (47.1) |  | 57 | (46.0) |  | 89 | (46.4) |
| Vascular dementia |  | 6 | (8.8) |  | 14 | (11.3) |  | 20 | (10.4) |
| Lewy body dementia |  | 1 | (1.5) |  | 6 | (4.8) |  | 7 | (3.6) |
| Frontotemporal dementia |  | 2 | (2.9) |  | 3 | (2.4) |  | 5 | (2.6) |
| Other |  | 18 | (26.5) |  | 38 | (30.6) |  | 56 | (29.2) |
| Unable to specify |  | 9 | (13.2) |  | 6 | (4.8) |  | 15 | (7.8) |

**Table 2S: Carer characteristics by arm for those in the extension study**

|  | **Randomised group** | | | | | | | | | |
| --- | --- | --- | --- | --- | --- | --- | --- | --- | --- | --- |
|  | **Routine care**  **N=68** | | | **NIDUS-Family intervention N=124** | | | | **Total**  **N=192** | | |
| **Carer Age (years), mean (SD)** |  | 63.7 | (10.6) |  | 61.4 | (11.5) |  | | 62.2 | (11.2) |
| **Carer Ethnicity, n(%)** |  |  |  |  |  |  |  | |  |  |
| White |  | 54 | (79.4) |  | 93 | (75.0) |  | | 147 | (76.6) |
| White other |  | 8 | (11.8) |  | 18 | (14.5) |  | | 26 | (13.5) |
| Mixed |  | 1 | (1.5) |  | 0 | (0.0) |  | | 1 | (0.5) |
| Asian |  | 1 | (1.5) |  | 6 | (4.8) |  | | 7 | (3.6) |
| Black |  | 1 | (1.5) |  | 5 | (4.0) |  | | 6 | (3.1) |
| Other |  | 3 | (4.4) |  | 2 | (1.6) |  | | 5 | (2.6) |
| **Carer First language, n(%)** |  |  |  |  |  |  |  | |  |  |
| English |  | 63 | (92.6) |  | 115 | (92.7) |  | | 178 | (92.7) |
| Other |  | 5 | (7.4) |  | 9 | (7.3) |  | | 14 | (7.3) |
| **Carer Gender, n(%)** |  |  |  |  |  |  |  | |  |  |
| Male |  | 25 | (36.8) |  | 29 | (23.4) |  | | 54 | (28.1) |
| Female |  | 43 | (63.2) |  | 95 | (76.6) |  | | 138 | (71.9) |
| **Carer Marital status, n(%)** |  |  |  |  |  |  |  | |  |  |
| Married/civil partnership |  | 54 | (79.4) |  | 94 | (75.8) |  | | 148 | (77.1) |
| Divorced |  | 4 | (5.9) |  | 1 | (0.8) |  | | 5 | (2.6) |
| Single |  | 3 | (4.4) |  | 16 | (12.9) |  | | 19 | (9.9) |
| Co-habiting |  | 4 | (5.9) |  | 10 | (8.1) |  | | 14 | (7.3) |
| Widowed |  | 3 | (4.4) |  | 1 | (0.8) |  | | 4 | (2.1) |
| Other |  | 0 | (0.0) |  | 2 | (1.6) |  | | 2 | (1.0) |
| **Carer Education, n(%)** |  |  |  |  |  |  |  | |  |  |
| Higher degree |  | 13 | (19.1) |  | 25 | (20.2) |  | | 38 | (19.8) |
| Degree |  | 21 | (30.9) |  | 40 | (32.3) |  | | 61 | (31.8) |
| A level (or equivalent) |  | 9 | (13.2) |  | 19 | (15.3) |  | | 28 | (14.6) |
| HNC/HND (or equivalent) |  | 5 | (7.4) |  | 4 | (3.2) |  | | 9 | (4.7) |
| NVQ (or equivalent) |  | 3 | (4.4) |  | 8 | (6.5) |  | | 11 | (5.7) |
| GSCE (or equivalent) |  | 11 | (16.2) |  | 18 | (14.5) |  | | 29 | (15.1) |
| School Leaving Certificate |  | 5 | (7.4) |  | 6 | (4.8) |  | | 11 | (5.7) |
| no formal qualifications |  | 1 | (1.5) |  | 4 | (3.2) |  | | 5 | (2.6) |
| **Relationship to care recipient, n(%)** |  |  |  |  |  |  |  | |  |  |
| Spouse/partner |  | 37 | (54.4) |  | 56 | (45.2) |  | | 93 | (48.4) |
| Child |  | 30 | (44.1) |  | 60 | (48.4) |  | | 90 | (46.9) |
| Friend |  | 0 | (0.0) |  | 1 | (0.8) |  | | 1 | (0.5) |
| Other |  | 1 | (1.5) |  | 7 | (5.6) |  | | 8 | (4.2) |
| **Carer Living situation, n(%)** |  |  |  |  |  |  |  | |  |  |
| Live alone |  | 2 | (2.9) |  | 6 | (4.8) |  | | 8 | (4.2) |
| Live with partner/spouse |  | 49 | (72.1) |  | 87 | (70.2) |  | | 136 | (70.8) |
| Live with flat/housemates |  | 0 | (0.0) |  | 1 | (0.8) |  | | 1 | (0.5) |
| Live with parent(s) |  | 7 | (10.3) |  | 9 | (7.3) |  | | 16 | (8.3) |
| Live with children |  | 2 | (2.9) |  | 3 | (2.4) |  | | 5 | (2.6) |
| Other |  | 8 | (11.8) |  | 18 | (14.5) |  | | 26 | (13.5) |
| **Carer Accommodation, n(%)** |  |  |  |  |  |  |  | |  |  |
| Council rented |  | 4 | (5.9) |  | 6 | (4.8) |  | | 10 | (5.2) |
| Housing association rented |  | 4 | (5.9) |  | 4 | (3.2) |  | | 8 | (4.2) |
| Private rented |  | 3 | (4.4) |  | 7 | (5.6) |  | | 10 | (5.2) |
| Owner-occupied |  | 57 | (83.8) |  | 103 | (83.1) |  | | 160 | (83.3) |
| Other |  | 0 | (0.0) |  | 4 | (3.2) |  | | 4 | (2.1) |

**Table 3S: Baseline characteristics for people living with dementia who were not in the extension study, by arm**

|  | Randomised group | | | | | | | | |
| --- | --- | --- | --- | --- | --- | --- | --- | --- | --- |
|  | Routine care  N=30 | | | NIDUS-Family intervention  N=80 | | | Total  N=110 | | |
| **Age (years) , mean (SD)** |  | 81.4 | (9.3) |  | 81.0 | (7.1) |  | 81.1 | (7.7) |
| **Ethnicity, n(%)** |  |  |  |  |  |  |  |  |  |
| White |  | 20 | (66.7) |  | 63 | (78.8) |  | 83 | (75.5) |
| White other |  | 5 | (16.7) |  | 4 | (5.0) |  | 9 | (8.2) |
| Mixed |  | 0 | (0.0) |  | 2 | (2.5) |  | 2 | (1.8) |
| Asian |  | 4 | (13.3) |  | 6 | (7.5) |  | 10 | (9.1) |
| Black |  | 1 | (3.3) |  | 5 | (6.2) |  | 6 | (5.5) |
| **First language, n(%)** |  |  |  |  |  |  |  |  |  |
| English |  | 24 | (80.0) |  | 71 | (88.8) |  | 95 | (86.4) |
| Other |  | 6 | (20.0) |  | 9 | (11.2) |  | 15 | (13.6) |
| **Gender, n(%)** |  |  |  |  |  |  |  |  |  |
| Male |  | 10 | (33.3) |  | 38 | (47.5) |  | 48 | (43.6) |
| Female |  | 20 | (66.7) |  | 42 | (52.5) |  | 62 | (56.4) |
| **Marital status, n(%)** |  |  |  |  |  |  |  |  |  |
| Married/civil partnership |  | 16 | (53.3) |  | 48 | (60.0) |  | 64 | (58.2) |
| Divorced |  | 2 | (6.7) |  | 6 | (7.5) |  | 8 | (7.3) |
| Single |  | 0 | (0.0) |  | 2 | (2.5) |  | 2 | (1.8) |
| Co-habiting |  | 0 | (0.0) |  | 1 | (1.2) |  | 1 | (0.9) |
| Widowed |  | 12 | (40.0) |  | 23 | (28.7) |  | 35 | (31.8) |
| **Education, n(%) (n=107)** |  |  |  |  |  |  |  |  |  |
| Higher degree |  | 2 | (6.7) |  | 11 | (14.3) |  | 13 | (12.1) |
| Degree |  | 6 | (20.0) |  | 15 | (19.5) |  | 21 | (19.6) |
| A level (or equivalent) |  | 2 | (6.7) |  | 5 | (6.5) |  | 7 | (6.5) |
| HNC/HND (or equivalent) |  | 1 | (3.3) |  | 4 | (5.2) |  | 5 | (4.7) |
| NVQ (or equivalent) |  | 1 | (3.3) |  | 1 | (1.3) |  | 2 | (1.9) |
| GCSE (or equivalent) |  | 5 | (16.7) |  | 8 | (10.4) |  | 13 | (12.1) |
| School Leaving Certificate |  | 7 | (23.3) |  | 18 | (23.4) |  | 25 | (23.4) |
| no formal qualifications |  | 6 | (20.0) |  | 15 | (19.5) |  | 21 | (19.6) |
| **Living situation, n(%)** |  |  |  |  |  |  |  |  |  |
| Live alone |  | 6 | (20.0) |  | 20 | (25.0) |  | 26 | (23.6) |
| Live with partner/spouse |  | 14 | (46.7) |  | 45 | (56.2) |  | 59 | (53.6) |
| Live with children |  | 7 | (23.3) |  | 11 | (13.8) |  | 18 | (16.4) |
| Other |  | 3 | (10.0) |  | 4 | (5.0) |  | 7 | (6.4) |
| **Accommodation, n(%)** |  |  |  |  |  |  |  |  |  |
| Council rented |  | 1 | (3.3) |  | 7 | (8.8) |  | 8 | (7.3) |
| Housing association rented |  | 1 | (3.3) |  | 3 | (3.8) |  | 4 | (3.6) |
| Private rented |  | 0 | (0.0) |  | 5 | (6.2) |  | 5 | (4.5) |
| Owner-occupied |  | 27 | (90.0) |  | 61 | (76.2) |  | 88 | (80.0) |
| Other |  | 1 | (3.3) |  | 4 | (5.0) |  | 5 | (4.5) |
| **Dementia diagnosis, n(%)** |  |  |  |  |  |  |  |  |  |
| Alzheimer’s Disease |  | 12 | (40.0) |  | 38 | (47.5) |  | 50 | (45.5) |
| Vascular dementia |  | 4 | (13.3) |  | 14 | (17.5) |  | 18 | (16.4) |
| Lewy body dementia |  | 2 | (6.7) |  | 1 | (1.2) |  | 3 | (2.7) |
| Frontotemporal dementia |  | 0 | (0.0) |  | 3 | (3.8) |  | 3 | (2.7) |
| Other |  | 8 | (26.7) |  | 20 | (25.0) |  | 28 | (25.5) |
| Unable to specify |  | 4 | (13.3) |  | 4 | (5.0) |  | 8 | (7.3) |

**Table 4S:** **Baseline characteristics for carers who were not in the extension study, by arm**

|  | N=30  Routine care | | NIDUS-Family intervention  N=80 | | Total  N=110 | |
| --- | --- | --- | --- | --- | --- | --- |
| **Carer Age (years), mean (SD)** | 64.7 | (13.4) | 65.7 | (14.5) | 65.4 | (14.2) |
| **Carer Ethnicity, n(%)** |  |  |  |  |  |  |
| White | 21 | (70.0) | 64 | (80.0) | 85 | (77.3) |
| White other | 3 | (10.0) | 5 | (6.2) | 8 | (7.3) |
| Mixed | 1 | (3.3) | 2 | (2.5) | 3 | (2.7) |
| Asian | 4 | (13.3) | 5 | (6.2) | 9 | (8.2) |
| Black | 1 | (3.3) | 4 | (5.0) | 5 | (4.5) |
| **Carer First language, n(%)** |  |  |  |  |  |  |
| English | 25 | (83.3) | 74 | (92.5) | 99 | (90.0) |
| Other | 5 | (16.7) | 6 | (7.5) | 11 | (10.0) |
| **Carer Gender, n(%)** |  |  |  |  |  |  |
| Male | 13 | (43.3) | 23 | (28.7) | 36 | (32.7) |
| Female | 17 | (56.7) | 57 | (71.2) | 74 | (67.3) |
| **Carer Marital status, n(%)** |  |  |  |  |  |  |
| Married/civil partnership | 23 | (76.7) | 62 | (77.5) | 85 | (77.3) |
| Divorced | 1 | (3.3) | 5 | (6.2) | 6 | (5.5) |
| Single | 6 | (20.0) | 8 | (10.0) | 14 | (12.7) |
| Co-habiting | 0 | (0.0) | 4 | (5.0) | 4 | (3.6) |
| Widowed | 0 | (0.0) | 1 | (1.2) | 1 | (0.9) |
| **Carer Education, n(%)** |  |  |  |  |  |  |
| Higher degree | 5 | (16.7) | 12 | (15.0) | 17 | (15.5) |
| Degree | 9 | (30.0) | 27 | (33.8) | 36 | (32.7) |
| A level (or equivalent) | 2 | (6.7) | 12 | (15.0) | 14 | (12.7) |
| HNC/HND (or equivalent) | 5 | (16.7) | 4 | (5.0) | 9 | (8.2) |
| NVQ (or equivalent) | 1 | (3.3) | 8 | (10.0) | 9 | (8.2) |
| GSCE (or equivalent) | 5 | (16.7) | 8 | (10.0) | 13 | (11.8) |
| School Leaving Certificate | 1 | (3.3) | 1 | (1.2) | 2 | (1.8) |
| no formal qualifications | 2 | (6.7) | 8 | (10.0) | 10 | (9.1) |
| **Relationship to care recipient, n(%)** |  |  |  |  |  |  |
| Spouse/partner | 14 | (46.7) | 46 | (57.5) | 60 | (54.5) |
| Child | 16 | (53.3) | 31 | (38.8) | 47 | (42.7) |
| Other | 0 | (0.0) | 3 | (3.8) | 3 | (2.7) |
| **Carer Living situation, n(%)** |  |  |  |  |  |  |
| Live alone | 2 | (6.7) | 3 | (3.8) | 5 | (4.5) |
| Live with partner/spouse | 20 | (66.7) | 61 | (76.2) | 81 | (73.6) |
| Live with flat/housemates | 0 | (0.0) | 1 | (1.2) | 1 | (0.9) |
| Live with parent(s) | 6 | (20.0) | 5 | (6.2) | 11 | (10.0) |
| Live with children | 0 | (0.0) | 5 | (6.2) | 5 | (4.5) |
| Other | 2 | (6.7) | 5 | (6.2) | 7 | (6.4) |
| **Carer Accommodation, n (%)** |  |  |  |  |  |  |
| Council rented | 1 | (3.3) | 4 | (5.0) | 5 | (4.5) |
| Housing association rented | 0 | (0.0) | 2 | (2.5) | 2 | (1.8) |
| Private rented | 0 | (0.0) | 6 | (7.5) | 6 | (5.5) |
| Owner-occupied | 28 | (93.3) | 66 | (82.5) | 94 | (85.5) |
| Other | 1 | (3.3) | 2 | (2.5) | 3 | (2.7) |

**Table 5S: Pattern mixture model analyses at 18 and 24 months (n=300) [δ are values subtracted from the MAR imputed GAS scores in each arm (from multiple imputation) before refitting regression models]**

| 18 month results | | | | | | | | | |
| --- | --- | --- | --- | --- | --- | --- | --- | --- | --- |
|  | NIDUS-Family intervention arm | | | | | | | | |
| Routine care arm |  | $\delta=0$ | | $\delta=-4$ | | $\delta=-8$ | | | $\delta=-12$ |
|  | $\delta=0$ | 12.43  [8.04, 16.82] | | 10.44  [6.03, 14.85] | | 8.45  [3.98, 12.92] | | | 6.45  [1.89, 11.02] |
|  | $\delta=-4$ | 14.04  [9.63, 18.44] | | 12.04  [7.62, 16.47] | | | 10.05  [5.56, 14.54] | | 8.06  [3.48, 12.64] |
|  | $\delta=-8$ | 15.64  [11.20, 20.08] | | 13.65  [9.19, 18.11] | | | 11.65  [7.14, 16.17] | | 9.66  [5.05, 14.27] |
|  | $\delta=-12$ | 17.24  [12.76, 21.73] | | 15.25  [10.74, 19.76] | | | 13.26  [8.69, 17.82] | | 11.27  [6.61, 15.92] |
| 24 month results | | | | | | | | | |
|  | NIDUS-Family intervention arm | | | | | | | | |
| Routine care arm |  | $\delta=0$ | $\delta=-4$ | | $\delta=-8$ | | | $\delta=-12$ | |
|  | $\delta=0$ | 9.27  [4.49, 14.06] | 6.97  [2.16, 11.78] | | 4.67  [-0.19, 9.52] | | | 2.36  [-2.58, 7.30] | |
|  | $\delta=-4$ | 11.13  [6.32, 15.94] | 8.83  [4.00, 13.65] | | | 6.52  [1.64, 11.40] | | | 4.22  [-0.74, 9.17] |
|  | $\delta=-8$ | 12.98  [8.14, 17.83] | 10.68  [5.82, 15.54] | | | 8.38  [3.47, 13.29] | | | 6.07  [1.08, 11.06] |
|  | $\delta=-12$ | 14.84  [9.95, 19.73] | 12.53  [7.63, 17.44] | | | 10.23  [5.27, 15.19] | | | 7.93  [2.89, 12.96] |

**Table 6S: Treatment effect estimates from mixed model analyses for primary and secondary outcomes**

| **Time point** | **Adjusted difference in means* (NIDUS – Routine care)** | **95% Confidence Interval** |
| --- | --- | --- |
| **Carer-rated GAS (primary outcome)** | | |
| **Main analysis (n=277)** | | |
| 6 months | 6.77 | [2.21, 11.34] |
| 12 months | 9.58 | [4.96, 14.20] |
| 18 months | 11.78 | [6.64, 16.93] |
| 24 months | 8.67 | [3.31, 14.02] |
| **Multiple imputation (n=300)** | | |
| 18 months | 12.43 | [8.04, 16.82] |
| 24 months | 9.27 | [4.49, 14.06] |
| **Worst-case sensitivity analysis (n=278)** | | |
| 18 months | 11.80 | [7.65, 15.96] |
| 24 months | 7.98 | [3.75, 12.20] |
| **Researcher-rated GAS (n=277)** | | |
| 6 months | 6.33 | [3.05, 9.60] |
| 12 months | 9.66 | [6.32, 13.00] |
| 18 months | 11.99 | [8.09, 15.90] |
| 24 months | 8.94 | [4.81, 13.06] |
| **DEMQOL-proxy (n=301)** | | |
| 6 months | -0.20 | [-3.07, 2.67] |
| 12 months | 2.68 | [-0.45, 5.80] |
| 18 months | 2.36 | [-1.59, 6.30] |

n = number of dyads in the model

Interaction term between arm and study visit is significant in the models for researcher-ratedGAS, Carer-rated GAS and Demqol-proxy (P-GAS p <0.001, C-GAS p < 0.001, Demqol-proxy p = 0.003).

*All models adjusted for study site and time-point with random effects to account for repeated measurements and facilitator clustering in the intervention arm. Interactions between randomised group and time were included in models to allow estimation of treatment effects at each follow up point. Models for DEMQOL-proxy included adjustment for baseline score.

**Table 7S: Summary of known deaths and permanent moves to care home occurring during each follow up period up to 24 months**

|  | 6 Months | | | 12 Months | | | 18 Months | | | 24 Months | | | Overall | | |
| --- | --- | --- | --- | --- | --- | --- | --- | --- | --- | --- | --- | --- | --- | --- | --- |
|  | N | n | (%) | N | n | (%) | N | n | (%) | N | n | (%) | N | n | (%) |
| Died, n(%) |  |  |  |  |  |  |  |  |  |  |  |  |  |  |  |
| Routine care | 98 | 3 | (3.1) | 84 | 6 | (7.1) | 63 | 4 | (6.3) | 55 | 0 | (0.0) | 98 | 13 | (13.3) |
| NIDUS-Family intervention | 204 | 6 | (2.9) | 170 | 6 | (3.5) | 114 | 8 | (7.0) | 92 | 5 | (5.4) | 204 | 25 | (12.3) |
| Move to care home, n(%) |  |  |  |  |  |  |  |  |  |  |  |  |  |  |  |
| Routine care | 98 | 2 | (2.0) | 84 | 1 | (1.2) | 63 | 8 | (12.7) | 55 | 4 | (7.3) | 98 | 15 | (15.3) |
| NIDUS-Family intervention | 204 | 2 | (1.0) | 170 | 5 | (2.9) | 114 | 12 | (10.5) | 92 | 6 | (6.5) | 204 | 25 | (12.2) |
| Transitioned (died or care home move) |  |  |  |  |  |  |  |  |  |  |  |  |  |  |  |
| Routine care | 98 | 5 | (5.1) | 84 | 7 | (8.3) | 63 | 11 | (17.5) | 55 | 4 | (7.3) | 98 | 27 | (27.6) |
| NIDUS-Family intervention | 204 | 8 | (3.9) | 170 | 11 | (6.5) | 114 | 19 | (16.7) | 92 | 10 | (10.9) | 204 | 48 | (23.5) |

**Table 8S: Frequency of reasons for irrelevance at 18 and 24 months**

| **Reason category** | | **18 months (N)** | **24 months (N)** | **Total (N)** |
| --- | --- | --- | --- | --- |
| Goal no longer felt realistic due to decline of person with dementia’s health | | 12 | 30 | 42 |
| Client died | | 30 | 4 | 34 |
| Client moved to care home | | 15 | 9 | 24 |
| Goal was achieved | Happy with the current outcomes | 3 | 8 | 11 |
|  | Alternative solutions found | 4 | 4 | 8 |
| Goal was no longer a priority | | 5 | 14 | 19 |
| No change observed | | 4 | 1 | 5 |
| External stressors and circumstances | | 0 | 3 | 3 |

**Table 9S: Example quotes from content analysis of reasons why goals no longer felt relevant**

| **Baseline goal area** | **Reason category** | **Reason no longer relevant provided** |
| --- | --- | --- |
| Interacting with others | **Goal was achieved (Dyad happy with the current outcome)** | Happy with level of activity, since 18m follow-up |
| Self-care (walking) | **Goal was achieved (Alternative solutions found)** | Not able to walk but being mobile in alternative way via scooter |
| Carer break | **Client died** | PLWD has died but I months prior to death FC had more time to self due to the additional help in place, enabled PWL to remain at home |
| Low mood | **Client moved to care home** | PLWD moved to care home, but mood appears much better |
| Self-care (physical complaints) | **Goal no longer felt realistic due to decline of person with dementia’s health** | Difficult now due to mobility problems being barrier to any physical activity |
| Self-care (physical complaints) | **Goal was no longer a priority** | No longer and important goal to the PLWD due to acceptance of decline |
| Memory of recent events (trouble with remembering to take their medications) | **External stressors and circumstances** | If FC is not in to give PLWD medication, she must arrange for someone else to come in to do so |
| Self-care (Increase fluid intake) | **No change** | PLWD has been this way all her life - all are aware this is an issue unlikely to change. |

**Figure 1S: Kaplan-Meier Survival Curve for Time to Death**

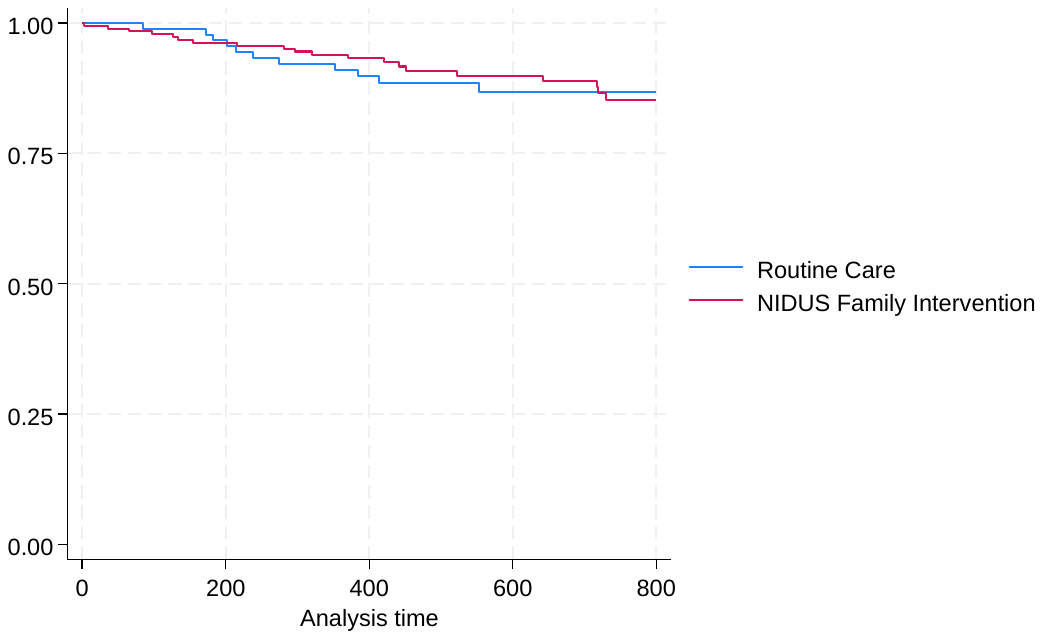

**Figure 2S: Kaplan-Meier Survival Curve for Time to Transition to care home (permanent move)**
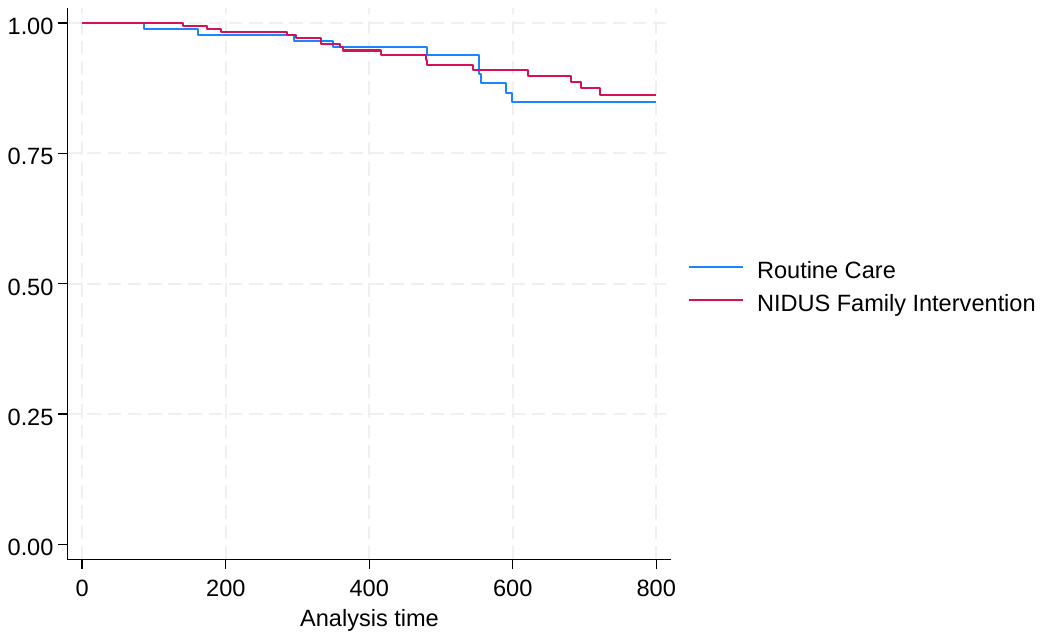
